# Kynurenines Increase MRS Metabolites in Basal Ganglia and Decrease Resting State Connectivity in Frontostriatal Reward Circuitry in Depression

**DOI:** 10.1101/2021.05.16.21257295

**Authors:** Xiangchuan Chen, Diana J. Beltran, Valeriya D Tsygankova, Bobbi J. Woolwine, Trusharth Patel, Wendy Baer, Jennifer C. Felger, Andrew H. Miller, Ebrahim Haroon

**Affiliations:** Emory Behavioral Immunology Program; Department of Psychiatry and Behavioral Sciences; Department of Anesthesiology, Emory University School of Medicine Atlanta, GA 30322

## Abstract

Inflammation is associated with depressive symptoms including anhedonia in patients with major depression. Nevertheless, the mechanisms by which peripheral inflammatory signals are communicated to the brain to influence central nervous system (CNS) function has yet to be fully elucidated. Based on laboratory animal studies, molecules of the kynurenine pathway (KP), which is activated by inflammation, can readily enter the brain, and generate metabolites that can alter neuronal and glial function, leading to behavioral changes. We therefore examined the relationship between KP metabolites in the plasma and cerebrospinal fluid (CSF) and brain chemistry and neural network function using multi-modal neuroimaging in 49 unmedicated, depressed subjects. CNS measures included 1) biochemical markers of glial dysfunction including glutamate (Glu) and myo-inositol (mI) in the left basal ganglia (LBG) using magnetic resonance spectroscopy (MRS); 2) local activity coherence (regional homogeneity, ReHo) and functional connectivity using resting-state functional magnetic resonance imaging; and 3) anhedonia from the Inventory for Depressive Symptoms-Self Reported. Plasma quinolinic acid (QA) was associated with increases and kynurenic acid (KYNA) and KYNA/QA with decreases in LBG Glu. Plasma kynurenine/tryptophan and CSF 3-hydroxy kynurenine (3HK) were associated with increases in LBG mI. Plasma and CSF KP were associated with decreases in ReHo in LBG and dorsomedial prefrontal cortex (DMPFC), and impaired functional connectivity between these two brain regions. DMPFC-BG connectivity mediated the effect of plasma and CSF KP metabolites on anhedonia. These findings highlight the contribution of KP metabolites to glial and neuronal dysfunction and ultimately behavior in depression.

## Introduction

Previous data has suggested that one mechanism by which inflammation can influence behavior is through effects on brain glutamate (Glu). Chronic administration of the inflammatory cytokine interferon (IFN)-alpha in patients with hepatitis C led to increased Glu in basal ganglia (BG) regions and anterior cingulate cortex (ACC) measured using magnetic resonance spectroscopy (MRS)^1,2^. Similar increases in BG Glu have also been reported in patients with major depression (MD) with increased endogenous inflammation, indexed by the acute phase reactant c-reactive protein (CRP)^3^. In each case, increased Glu in these brain regions was associated with increases in the overall severity of depressive symptoms and symptoms of anhedonia, amotivation, and psychomotor slowing^1–3^. In addition, greater CRP concentrations in the cerebrospinal fluid (CSF) of MD patients were associated with increases in myo-inositol (mI), a marker of astroglial distress in the BG, suggesting that the effects of inflammation upon Glu may reflect alterations in astrocyte function induced by inflammation and oxidative stress^3,4^.

Relevant to the impact of inflammation on Glu and astrocytic function, there has been an increasing interest in the potential role of metabolites of the kynurenine pathway (KP). Inflammatory stimuli such as lipopolysaccharide (LPS) and Bacillus Calmette–Guérin (BCG) vaccine and inflammatory cytokines such as tumor necrosis factor (TNF) and IFN gamma activate the enzyme indoleamine 2,3 dioxygenase (IDO), which converts tryptophan (TRP) to kynurenine (KYN)^5–7^. Under homeostatic conditions, ~95% of TRP is converted into KYN, and its breakdown products culminating in the generation of nicotinamide adenine dinucleotide (NAD+), an important cellular energy source^7,8^. KYN is then transported into the brain, where it is metabolized either into 3-hydroxykynurenine (3HK), anthranilic acid (AA), 3-hydroxyanthranilic acid (3HAA), and quinolinic acid (QA) in microglia, tissue macrophages, and trafficking monocytes; or kynurenic acid (KYNA) in astrocytes, oligodendrocytes, and neurons^8–12^. KYN and 3HK gain access to the brain via the large neutral amino acid transporter (LAT-1) in the blood-brain barrier (BBB), leading to an excess of KP metabolism in the brain^12–14^. These data are consistent with findings in patients with MD, whereby path analyses have demonstrated that increased peripheral blood inflammatory markers are linked to increased peripheral blood KP metabolites that are in turn associated with increased KP metabolites in CSF^15^. Of note, in the context of chronic immune activation, astrocytes generate large quantities of KYN that can be converted into QA by adjacent macrophages and microglia^16^. KYN can also amplify local proinflammatory signaling via activation of the nod-like receptor protein (NLRP)-2 inflammasome complex in astrocytic cells^17^. Astrocytes are responsible for most Glu uptake in synaptic and nonsynaptic areas and, consequently, are the major regulators of Glu homeostasis^18^.

KP metabolites influence glutamatergic activity in several distinct ways^19^. QA is a well-known endogenous neurotoxin and gliotoxin^20^. Regarding the impact of KP metabolites on astrocytic regulation of Glu, QA directly promotes astrocytic Glu release, increases its cycling, and decreases its reuptake by astrocytic transporters^19,20^. 3HK also increases astrocytic Glu release via the induction of reactive oxygen species, leading to activation of the system xC-antiporter, which extrudes one molecule of Glu in exchange for a molecule of cysteine that is used in the synthesis of the antioxidant glutathione^21,22^. Another target for KP metabolites is the N-methyl-D-aspartate (NMDA)-glutamate receptor on glutamatergic neurons, with QA acting as an agonist and KYNA acting as an antagonist and allosteric modulator at the glycine site ^19,20,22^.

These effects of KP metabolites in conjunction with inflammation on astrocytes and glutamatergic receptor signaling can contribute to excessive Glu release and spillover into the extrasynaptic space^23,24^. The extrasynaptic diffusion of Glu leads to activation of extrasynaptic NMDA receptors^25–28^, ultimately influencing the concordance and synchrony of BOLD fluctuations between neighboring voxels in resting-state functional MRI (rsfMRI) as reflected by regional homogeneity (ReHo)^29^. Patterns of spontaneous brain oxygen level-dependent (BOLD) oscillatory activity in rsfMRI^30–37^ provide yet another opportunity to study the relationship between MRS metabolites and functional brain activity. Indeed, MD patients with concurrent increases in inflammation and CNS Glu exhibited ReHo decreases at the site of MRS acquisition and over a wider network of brain regions, particularly those involved in reward processing^29^. Impaired connectivity between regions with decreased ReHo was associated with greater depression, anhedonia, and psychomotor slowing^29^. Our previous data and those from other groups have consistently linked concurrent increases in inflammation and KP metabolism with depression, suicidality, anhedonia, and non-response^15,38–42^. These findings are also consistent with a rich literature linking the development of depression-like behavior in preclinical models (e.g., decreased sucrose preference, greater immobility on forced-swim or tail-suspension tests) following IDO-activation by inflammatory stimuli^7,12,43–46^. Finally, in laboratory animals, blocking KP activation through inhibition of IDO or inhibiting the transport of KYN into the brain using leucine or blocking of NMDA receptors with ketamine can reverse inflammation-induced depressive-like behavior^7,14,44,46^.

Nevertheless, no study has examined the associations of plasma and CSF KP metabolites with MRS and rsfMRI measures (Glu, mI, ReHo, and connectivity) in patients with MD. We, therefore, examined the impact of plasma and CSF KP on brain chemistry and function of BG using MRS and rsfMRI. We hypothesized that increased plasma/CSF KP metabolites would be associated with elevated MRS-signals of astroglial dysfunction (Glu and mI), reduced ReHo, and decreased functional connectivity in BG and other reward-related brain regions, resulting in greater severity of anhedonia. Identifying such a biomarker/neuroimaging-based biosignature could help identify a distinct subgroup of depressed patients with increased inflammation who may respond to KP-targeted pharmacotherapeutics.

## Materials and Methods

### Study Sample

We included unmedicated subjects between the ages of 21-65 years with a primary diagnosis of MD based on Structured Clinical Interview for Diagnostic and Statistical Manual-IV (SCID-IV)^47^ and a 17-Item Hamilton Rating Scale for Depression score (HAM-D) ≥18^48^. Subjects with active suicidal ideation (score >2 on item #3 on HAM-D) were excluded from study participation and referred for treatment. All qualifying patients were monitored for safety (e.g., suicidality, worsening) and referred for further management if necessary. We determined study eligibility based on history and physical examination, psychiatric evaluation, laboratory testing, and electrocardiography. Presence of Axis I psychiatric disorders (other than anxiety); substance abuse/dependence within the past 6-months (SCID-IV criteria and urine drug testing); unstable medical disorders (needing multiple visits or medication changes); intake of psychotropic medications of any kind (other than prn benzodiazepines, last drug intake > 72 hours prior to assessments); or intake of medications known to affect the immune system (e.g. anti-cytokine therapies, methotrexate, oral steroids, non-steroidal anti-inflammatory agents) were exclusionary. Patients with autoimmune or inflammatory disorders, chronic infections (hepatitis B or C and Human Immunodeficiency Virus) or a history of cancer (other than basal cell skin cancer) were excluded. No individual discontinued their psychotropic medications for the purposes of the study. The Institutional Review Board of Emory University approved the study, and all subjects provided written informed consent. Subjects presented herein represent a subset of patients recruited for an NIH-funded study on phenotyping depressed patients with increased inflammation and overlap with subjects that have appeared in previous reports (NCT01426997)^3,49^.

### Assessment of Depression Severity and Anhedonia

Standardized measures of depression severity were obtained using the 30-item Inventory of Depressive Symptoms - Self-Reported (IDS-SR) – a scale that has been extensively used in previous studies by our and other groups^3,49–51^. IDS-SR was completed in 1 sitting, and the patients were asked to choose the item response (0, 1, 2, or 3) that best described themselves over the past seven days. The scale was scored by adding 28 of 30 items to obtain a total score ranging from 0-84, excluding items #11-12 (Appetite increase/decrease) and #13-14 (Weight increase/decrease) - per instructions from the developers^51^. In addition, items with ambiguous ratings including #9 (‘Diurnal Variation of Mood’ with two subitems 9A and 9B), #10 (‘Quality of Mood’ with difficulty in differentiating depression versus grief), and #25 (Aches and Pains with difficulty in differentiating items from other somatic symptoms) were excluded based on patient feedback. Hierarchical clustering (**Results, Figure 3**) of the remaining 23 individual IDS-SR items indicated that four anhedonia items (#8: “Response to Good or Desired Events,” #17: “View of My Future,” #19: “General Interest Excluding Sex,” and #22: “Interest in Sex”) tightly clustered together. Anhedonia was determined as a factor score representing individual ratings on these four items.

**Figure 1:**
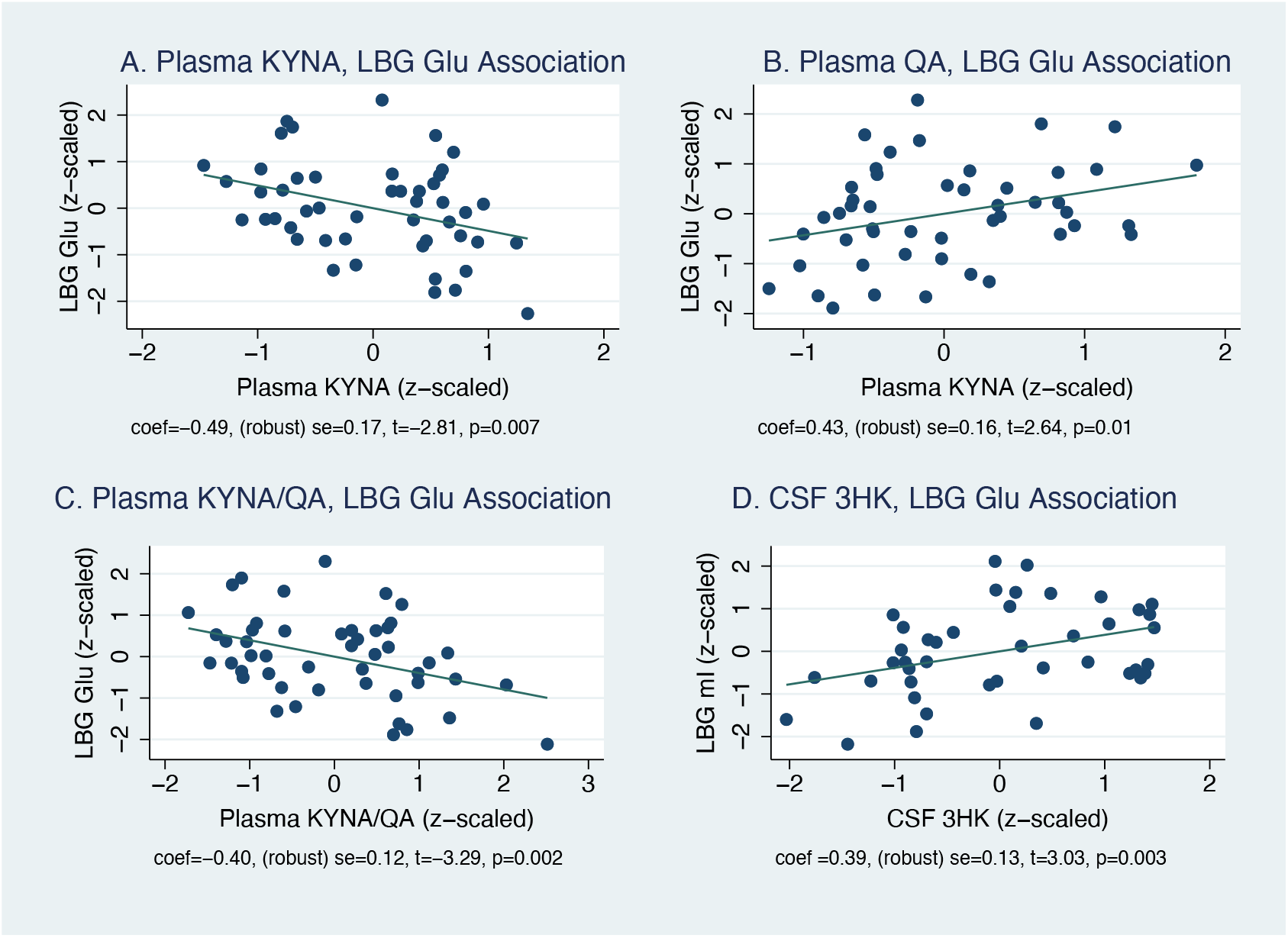
KP-MRS Associations. **Legend: Figure 1** represents the partial (A, B) and simple regression plots (C, D) demonstrating the association between kynurenine pathway (KP) metabolites on the x-axis and magnetic resonance spectroscopy (MRS)–based markers in the left basal ganglia on the y-axis. All values were normalized (using Box-Cox Power Transform) and z-scaled. These are simplified least squares models compared with the more complex lasso regression used in the text. **Figure 1A** represents the association between plasma kynurenic acid (KYNA) and LBG glutamate (LBG Glu), **Figure 2B** between plasma quinolinic acid (QA) and LBG glutamate (Glu), **Figure 1C** between plasma KYNA/QA ratio and LBG Glu, and **Figure 1D** between cerebrospinal fluid (CSF) 3-hydroxy kynurenine (3HK) and LBG myo-inositol (LBG mI). The individual coefficients and their significance values are indicated under each graphlet.

**Figure 2:**
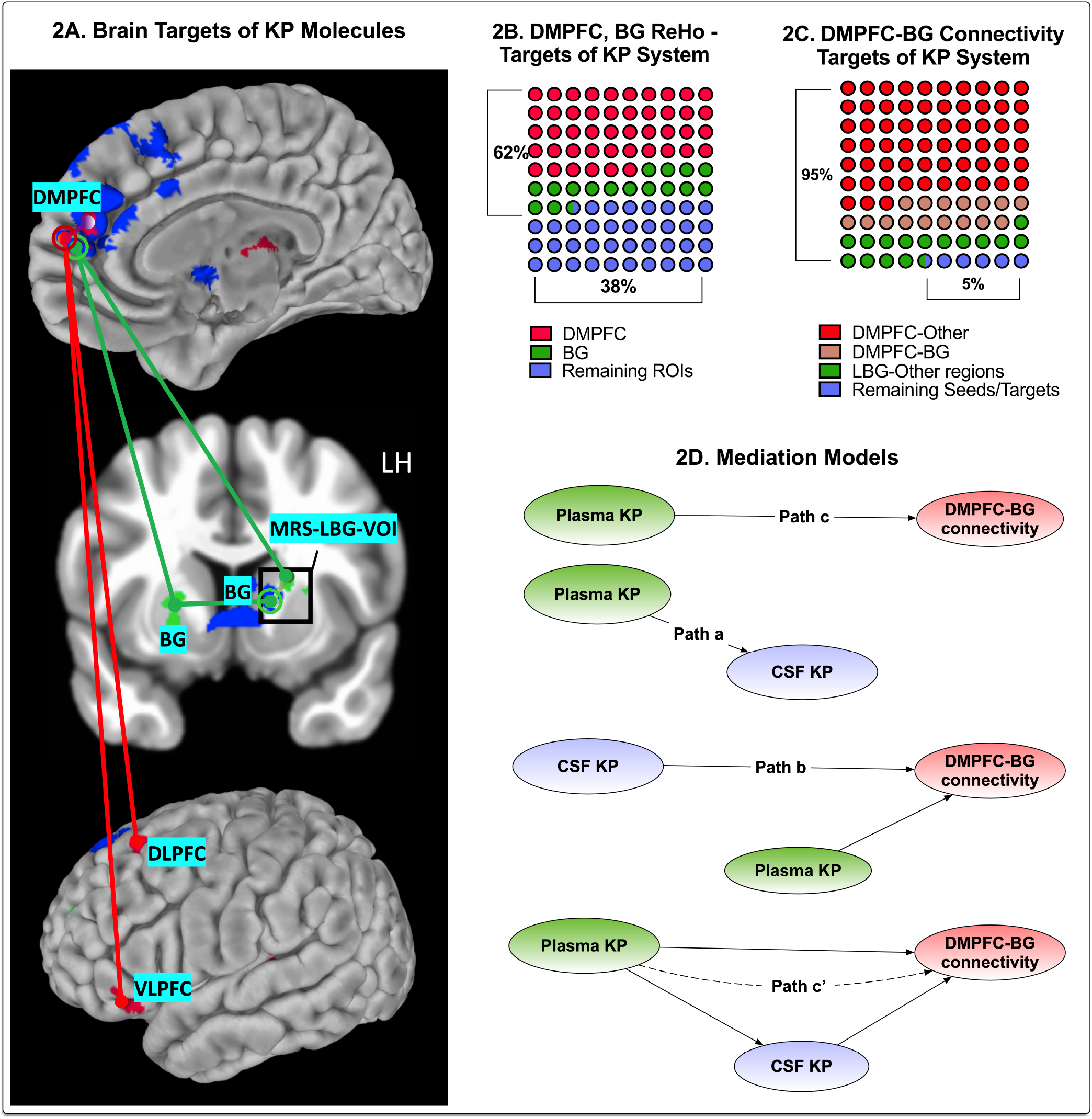
KP-Resting fMRI Associations. **Legend: Figure 2A** presents the primary resting-state functional magnetic resonance imaging (rsMRI) targets of kynurenine pathway (KP) metabolites identified using voxel-based analysis. The areas shaded in deep blue represent the regions of interest (ROI) that demonstrated altered Regional Homogeneity (ReHo) associated with plasma or cerebrospinal fluid (CSF) KP. Identification of ROIs was based on the correlation between kynurenine (KYN) or 3-hydroxy-kynurenine (3HK) in either the plasma or CSF. A detailed description of all ReHo seed-ROIs thus derived (total = 12 seeds) is presented in **Supplemental Information 4**. ROIs correlated with plasma KP are illustrated using red circles, and CSF KP-related ROIs are illustrated using green circles. These ROIs were also set as the seeds in seed-to-whole brain connectivity analysis. The red or green lines and the red or green uncircled dots represent the connectivity and targets, respectively. Connectivity z-scores correlated with plasma and CSF KP measures were derived after controlling for multiple comparisons (threshold = voxel p<0.001, cluster p<0.05). **Figure 2B** presents a 10×10 dot plot depicting the region-wise distribution of voxels (measured using voxel volume=1 cubic millimeter/voxel), demonstrating ReHo decreases. As is evident, 62% of voxels demonstrating decreased ReHo were located either in the dorsomedial prefrontal cortex (DMPFC), left (LBG), or right basal ganglia (RBG) regions, respectively. **Figure 2C** demonstrates a similar dot plot demonstrating the distribution of KP-correlated connectivity disruptions between ReHo seed regions and other brain regions. As is evident, 95% of whole-brain connectivity decreases were associated with DMPFC-BG regions. **Figure 2D** represents the overall plan of the path/mediation analyses. Plasma and CSF KP latent factors represent the individual contribution of kynurenine (KYN), 3-hydroxy kynurenine (3HK) to each of these factors. DMPFC-basal ganglia (BG) latent factor comprises four different connectivities (DMPFC-LBG, DMPFC-RBG, LBG-LBG, and LBG-RBG). **Path c** examines the relationship between plasma KP and DMPFC-BG connectivity, and **Path a** examines the association between plasma and CSF KP to each other. **Path b** examines if plasma KP impacts the association between CSF KP and DMPFC-BG connectivity. **Path c’** presents the model testing mediation by CSF KP of the relationship between plasma KP on DMPFC-BG connectivity.

**Figure 3:**
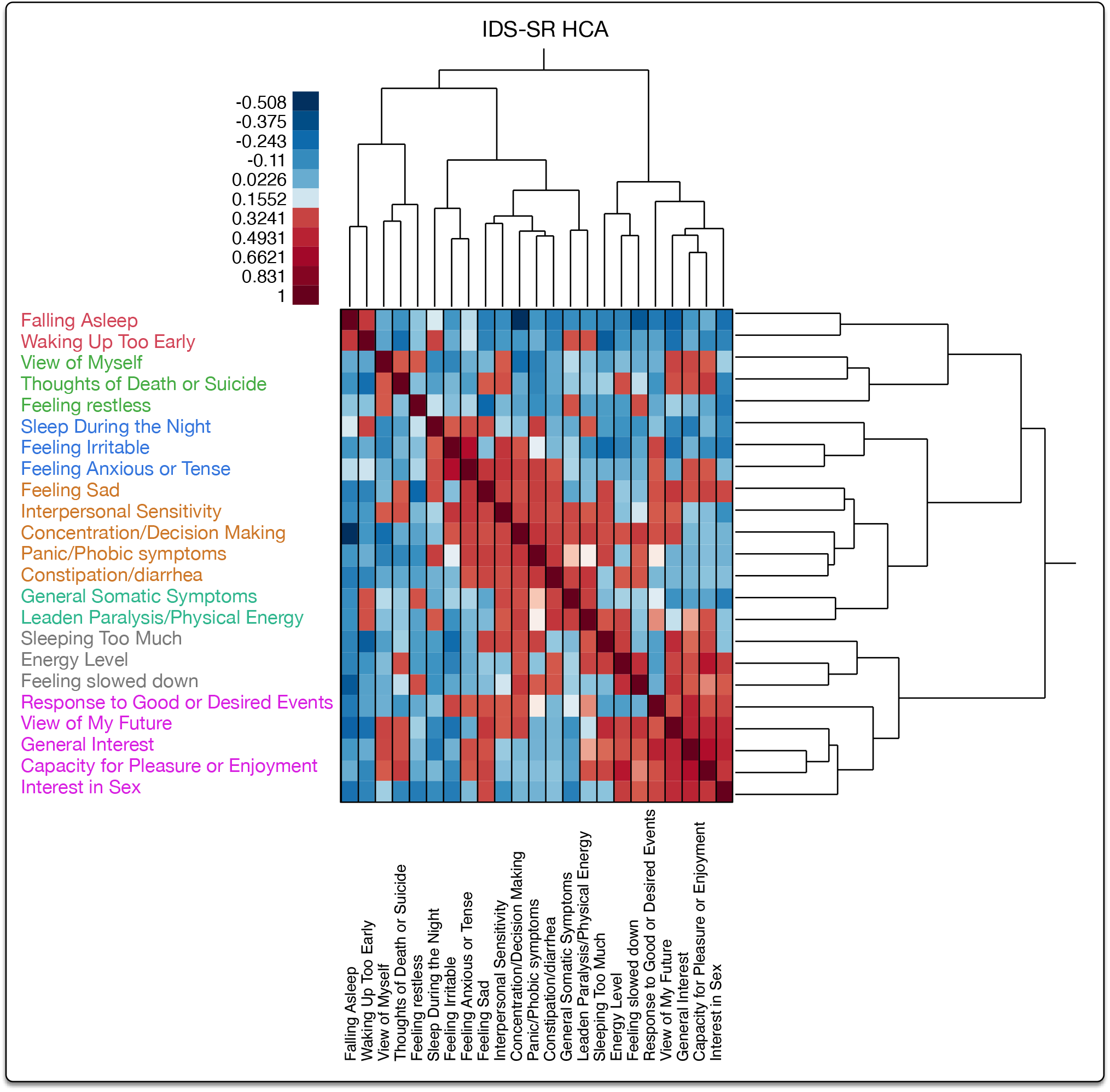
Anhedonia – HCA Clustering. **Legend**: **Figure 3** depicts the dendrogram from hierarchical clustering of individual items on Inventory for Depressive Symptoms-Self Reported (IDS-SR). After removing items with ambiguous and unreliable scoring on IDS-SR (details in text), the remaining 23 items were clustered using hierarchical clustering with Ward’s agglomerative method. As noted, anhedonia items on IDS-SR (items #8, 17, 18, 19, and 22) clustered tightly together and were combined into an ‘Anhedonia’ factor. Item #18: Capacity for Pleasure or Enjoyment was removed during redundancy analysis, as it failed to load adequately onto its latent factor.

### Blood Sampling and Assays

Blood sampling was conducted via an indwelling catheter between 0800-1000hrs, and all subjects sat quietly for 30 minutes before phlebotomy to minimize the impact of circadian variations and stress. The sampled blood was immediately centrifuged at 1000g for 10 minutes at 4°C. Plasma was then removed and frozen at −80°C for batched assays. Lumbar puncture (LP) was performed at the Emory General Clinical Research Center between 1100 and 1500hrs by an attending anesthesiologist one day after blood sampling, as described previously^15,52^. Approximately 10mL of CSF was collected for each subject after discarding the initial 1mL to avoid blood contamination. Samples were collected into chilled tubes, aliquoted into 1mL vials, and immediately frozen at −80°C until batched assays. **KP assays:** Concentrations of TRP, KYN, KYNA, 3HK, AA, 3HAA, and QA in plasma and the CSF were measured blind to diagnosis by Brains Online, LLC (Charles River, Inc). The plasma and CSF metabolite concentrations were estimated using high-performance liquid chromatography (HPLC) with tandem mass spectrometry (MS/MS) detection using standardized protocol per specifications provided by the vendor. The overall assay CV was <10% for all KP metabolites in the plasma and CSF (see **Supplementary Information 1** for assay details, and **Supplementary Table 1** for assay reliability). All measurements with unreliable quantitation were excluded. KYN/TRP and QA/KYNA ratios were computed for further analyses. **Immune assays:** Based on our previous work showing a strong relationship with KP metabolites in plasma and CSF, TNF and TNF receptor-type-2 (TNFR2) were measured in plasma and CSF based on specifications detailed in our previous studies^15^.

### MRS/MRI Analysis

MRI scans were acquired using a Siemens 3T Trim-Trio scanner (Siemens Medical Solutions, Malvern, PA, USA). Head motion was limited with foam restraints. T1 images were acquired using MPRAGE sequence (TR/TE/ TI=2300/3.02/1100 ms, flip angle (FA=8o°, voxel size= 1×1×1 mm^3^, matrix size=256×256). rsfMRI data were acquired using an EPI sequence (TR/TE=2950/30 ms, FA=90°, FOV=220 mm, matrix size=3.4×3.4×4.0 mm^3^, slice number=30, 150 repetitions). Participants were instructed to keep their eyes open and look at a fixed crosshair on the scanner’s projection screen. Single-voxel MRS (MRS-SVS) data were acquired using a PRESS sequence (TR/TE=3000/30 ms, FA=90°, sampling size = 1024, size of voxel of interest [VOI]=30×17 x17 mm^3^, 128 averages with water suppression, 4 with water unsuppressed scans). The VOI was placed in the LBG area (Figure 2A). The preprocessing steps are described in **Supplemental Information 2** and our previous publications^3,31^. The data were preprocessed by correcting signal spikes, slice-timing shift, and head motion, and removing motion and CSF signals with a general linear model (GLM). The residual time-series data were used to calculate ReHo, converted into the MNI space, and spatially smoothed (full-width half-maximum, FWHM=4 mm). KP-related brain regions of interest (ROIs) were identified by correlating the ReHo results with KP measures. The analysis was conducted using recommended cluster-wise multiple-test correction (voxel-level p<=0.001 and cluster-wise p<=0.05^53,54^. ReHo-KP marker correlations were limited to KP metabolites found to be significant in the KP-MRS analysis to avoid multiple comparisons. KP-related ReHo ROIs thus identified were set as seeds for the connectivity analysis. After further removal of the WM signals and temporal filtering (0.01–0.1 Hz), averaged time-series data was calculated for each ReHo ROI and then correlated with the time series data of individual voxels in the brain. Connectivity data from the CSF-KP ReHo ROIs were correlated with the CSF-KP measures, and those from the plasma-KP ReHo ROIs were correlated with the plasma-KP measures. Individual CSF and plasma KP measures were used independently to define both the seed (ReHo ROIs) and itheir target connectivity ROIs after applying the above-mentioned multiple correction threshold. Partial-volume correction of absolute quantitation of metabolite concentrations from MRS was performed based on the volumetric ratios of GM, WM, and CSF in the VOI, as described in our earlier publications^3,29^. See **Supplementary Information 2** for more details of MRI data acquisition and analysis.

### Statistical Analysis - Lasso-Inferential Model

Penalizing, cross-fit, partialing-out (double machine learning), lasso-inferential models robust to collinearity were used in Stata v16 (Stata, College Station, TX, USA) to examine the association between MRS markers and KP metabolites after controlling for age, sex, race (Caucasian vs. African American) and body mass index (BMI). Independent models for each of the MRS markers (Glu and mI) and KP metabolites from each compartment (plasma and CSF) were used after partialing out effects of covariate/control variables (age, sex, BMI, and race) per Frisch-Lovell-Waugh theorem. The analysis was geared to test the explanatory power of the primary variables of interest (primary model) and to examine the effect of adding or removing covariates upon these associations (auxiliary model). We conducted this analysis in two phases. The primary model used a plugin estimator consisting of k(10)-fold*resample(10) runs (total=100 runs), and the secondary (sensitivity) analysis used alternate estimators, including cross-validation (CV) and adaptive methods. Of note, noise factors induced by control covariates do not disrupt the primary (plugin) estimator than the auxiliary (cross-validated or adaptive) estimators.

### Partial Least Squares - Structural Equation Modeling

(PLS-SEM) was used to examine path effects and mediational associations in Smart PLS^55^. Biomarkers (KP, immune), seed-connectivities, and anhedonia items (of IDS-SR) were used as indicators to generate latent constructs (measurement model) that were further used to test hypotheses based on path associations (structural model). Moderation of these associations by covariates was tested using linear path models for continuously distributed variables (age, BMI) and using group comparison models for categorical covariates (sex, race). The precise details of the measurement and structural model are provided in **Supplemental Information 3**.

## Results

### Background Data

Forty-nine subjects with available KP and MRS measures were recruited, of whom 46 subjects with available plasma KP and MRS measures and 40 with KP and rsfMRI measures were included in the study. Details on missing information are provided in **Supplementary Table 1 and Supplementary Information 4. Table 1** provides details of the background demographic, clinical and biological data. None of the missing biomarker data were imputed as it was judged that the missingness was not random. One subject did not answer item #22 on IDS-SR (item on Libido), and the result was imputed using low-rank approximation after verifying it was missing completely at random. The subjects were in moderate to severely depressed range based on HAM-D Scores (mean±SD=23.2±3.2), aged between 21-64 (mean±SD=38.6±10.9 years), and obese (body mass index (BMI)=31.8±7.9 kg/m2). 69% of the subjects were women, and African Americans comprised 65% of the study population. The high percentage of women is a characteristic of most depressed subject groups, and the high representation of African Americans is consistent with the study location (Atlanta, GA). Several values of CSF KYNA, QA, 3HAA were discarded due to estimation difficulties by BrainsOnline, Inc. Hence only 4 CSF KP metabolites (TRP, KYN, 3HK, and AA) and one ratio (CSF KYN/TRP) were available for analysis.

**Table 1:**
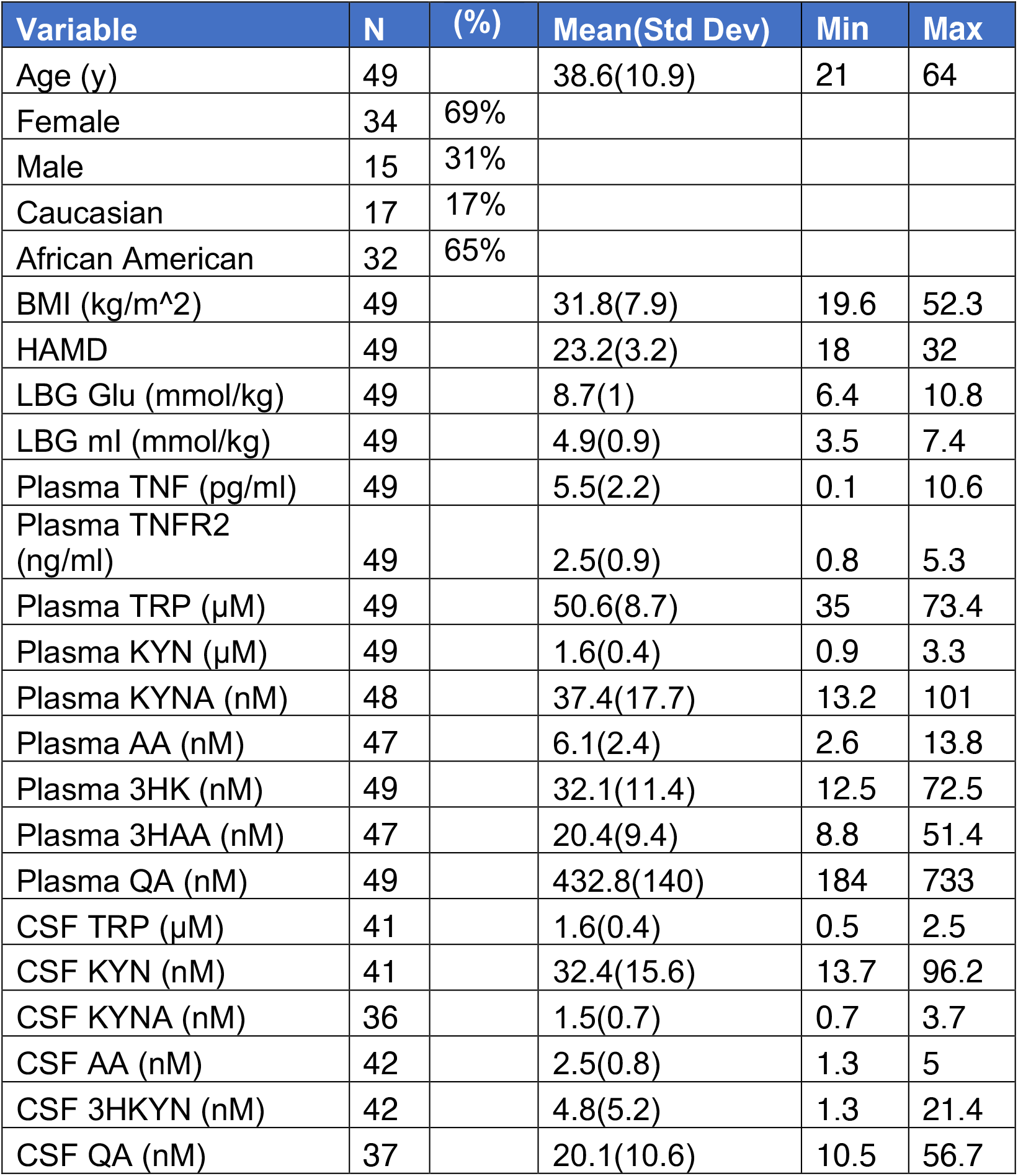
Demographic, Clinical and Biological Variables of the Sample.

### Plasma KYNA, QA, and KYNA/QA association with MRS Glu

Using the cross-fit, partialing out lasso, plasma QA (**Figure 1A**) was positively (Cf=0.53, 95%CI(0.07,0.99), z=2.41, p=0.024) and plasma KYNA (**Figure 1B**) was negatively (Cf=−0.40, 95%CI=(−0.74,-0.05), z=−2.25, p=0.02) associated with left BG (LBG) Glu. The associations survived multiple testing by K(10)-fold with 10-resamplings of each fold in the primary analysis and during the sensitivity analysis, indicating relative insensitivity to perturbation by covariate induced noise factors. None of the other plasma or CSF metabolites were significantly associated with LBG Glu. Plasma KYNA/QA ratio was negatively associated with LBG Glu (**Figure 1C**, Cf(95%CI)=−0.40(−0.63, −0.16), z=−3.27, p=0.001). The relationship between plasma KYNA/QA was robust (survived 100-fold resampling) and was not perturbed by adding or removing covariate/control variables during sensitivity analysis using CV and adaptive estimations.

### Plasma KYN/TRP and CSF 3HK associations with LBG mI

Plasma KYN/TRP ratio was positively associated with LBG mI (Cf(95%CI)=0.25(0.02, 0.48), z=2.17, p=0.03) only in the primary model. This association did not survive sensitivity analysis due to a strong interaction between mI and age. The potent oxidative stress-inducing molecule CSF 3HK (**Figure 1D**) was associated with LBG mI (Cf(95%CI)=0.45(0.19, 0.75), z=3.35, p=0.001) in the primary model after 100-fold repetitions. However, this association did not survive sensitivity analysis as age was independently associated with mI, while BMI impacted CSF 3HK. None of the other plasma or CSF KP makers or CSF KYN/TRP were associated with LBG mI.

### KP associations with rsfMRI measures (Figure 2, Supplemental Information 6)

Based on the above results, only KYN, 3HK, KYNA, QA, KYN/TRP, and KYNA/QA ratios in the plasma, and KYN and 3HK in the CSF were used for ReHo analysis. Plasma QA, KYNA and KYNA/QA yielded ReHo ROIs identical to those provided by KYN and 3HK (provided in **Supplementary Results, Supplementary Figures 2, 3**). Accordingly, ReHo data correlated with plasma KYN, plasma 3HK, CSF KYN, and CSF 3HK were used from all 40 subjects. ReHo ROI correlated with the above 4 markers were used as originating seeds to identify altered connectivity between these and other brain regions as a function of the four KP metabolites. After adjusting for multiple comparisons (voxel p<=0.001, cluster p<=0.05), the 4 KP measures were associated with decreased ReHo in a total of 12 ROIs. Besides, 19-decreased seed connectivities between these ROIs and other brain regions were also noted (**Figure 2A**). **Supplementary Information 5, Supplementary Tables3-4, Supplementary Figures 1** provide a full list of ReHo ROIs and seed connectivities related to the KP metabolites. In terms of voxel volume, ReHo decreases in the DMPFC, and BG together accounted for 62% of total brain wide ReHo decreases identified by all 4 KP metabolites (**Figure 2B**). Additionally, 90% of seed-connectivity decreases explained by the above 4 KP measures also involved either DMPFC or BG ROIs (**Figure 2C**).

### CSF-KP mediates the effects of Plasma KP on DMPFC-BG connectivity (Figure 2D)

PLS-SEM was used for path analysis using Baron and Kenny’s four-step approach^56^. The mediation model included three factors, i.e., Plasma KP- and CSF KP-factors (latent factors generated from KYN and 3HK from plasma and CSF, respectively), and DMPFC-BG connectivity factor correlated with metabolites used to derive CSF KP latent (including DMPFC-LBG, DMPFC-right (R)BG, LBG-LBG, and LBG-RBG). **Step 1 (path ‘c’)** examined if there is an effect to mediate. We examined if Plasma KP-latent (predictor variable) was associated with DMPFC-BG connectivity latent. All values reported below were obtained using 10000-bootstrapped repetitions. Plasma KP-factor was associated with DMPFC-BG-factor with a negative path (Cf(95%CI)=−0.56(−0.78, −0.37), t=3.36, p=0.001) with a strong effect size (r-sq=0.31, Cohen’s f-sq=0.45). **Step 2 (path ‘a’)** examined if the predictor variable was correlated with the mediator. We used CSF KP (mediator) as the criterion variable in the path model and Plasma KP as a predictor to estimate and test path ‘a’. In line with our previous reports^17^, plasma KP-factor was associated with CSF KP-factor via a positive direct path (Cf(95%CI)=0.67(0.45, 0.88), t=5.57, p<0.001). **Step 3 (path ‘b’)** examined if the mediator (CSF KP) affects the outcome variable (DMPFC-BG connectivity latent) after controlling for Plasma KP. We used DMPFC-BG connectivity as the criterion variable in the path model with Plasma KP and CSF KP as predictors. CSF KP was associated with DMPFC-BG connectivity latent via a strong negative effect (Cf(95%CI)=−0.67(−0.99, −0.43), t=4.82, p<0.001), after controlling for the effect of Plasma KP-latent. Of note, Plasma KP/DMPFC-BG connectivity association lost its previously noted significance in this step. **Step 4 (path c’)** tested the hypothesis that CSF KP completely mediates the effect of the Plasma KP/DMPFC-BG relationship. The effect of controlling for CSF KP (mediator variable) would diminish (partial mediation) or nullify (complete mediation) the Plasma KP/DMPFC-BG association. CSF KP-factor exerted a full mediation effect on the association between plasma KP and DMPFC-BG (path c’, Cf(95% CI)=−0.44(−0.89, −0.18), t=2.35, p=0.019). The full-mediation effect by CSF KP on the association between Plasma KP and DMPFC-BG connectivity had strong predictive power and replicability for both in-sample (r-sq difference=0.20, f-sq=0.41) and out-of-sample (blindfolding cross-validation test Q-sq difference=0.22, f-sq=0.34) metrics.

### Moderation of KP/Connectivity Associations by immune markers, BMI, and other covariates

Moderation of above path c’ by immune markers (TNF/TNFR2), BMI, age, sex, and race were examined. Plasma TNF/TNFR2 was associated with DMPFC-BG connectivity latent via a negative path (Cf(95%CI)=−0.29(−0.55, −0.11), t=2.52, p=0.01). Plasma KP->CSF KP significantly mediated this association with a strong effect size (mediation effect Cf(95%CI)=−0.25(−0.57, −0.09), t=2.02, p=0.043, f-sq=0.31). A similar negative indirect association between BMI and DMPFC-BG connectivity was noted (Cf(95%CI)=−0.16(−0.39, −0.05), t=2.10, p=0.03), which was similarly mediated by Plasma->CSF KP (Cf(95%CI)=−0.16(−0.33,-0.05), t=2.27, p=0.023). Age, sex, and race did not exert moderating effects on the above paths.

### DMPFC-BG connectivity/Anhedonia Association

Anhedonia latent was developed using loading from 4 items (#8, 17, 19, 22) of IDS-SR that clustered together during hierarchical clustering, as shown in Figure 3. Item #18: Capacity for Pleasure or Enjoyment was removed following redundancy analysis, as it failed to load adequately in its latent factor (loading coefficient <0.5). 3/19 connectivities correlated with plasma KYN (DMPFC-DMPFC, DMPFC-left ventrolateral prefrontal region, DMPFC-left dorsolateral prefrontal region), and 1/19 related to CSF 3HK (DMPFC-LBG) were selected based on their significant loadings on to the path model predicting Anhedonia Factor (loading coefficient >0.7). **Step 1 (Figure 4A)** examined the direct relationship between KP-associated functional connectivity and Anhedonia. A significant negative association was noted between the DMPFC-BG connectivity factor and the Anhedonia factor (Cf(95%CI)=−0.56(−0.77,-0.43), t=6.0, p<0.001), with a strong effect size (r-sq=0.36, Cohen’s f-sq= 0.45). **Step 2** involved combining all latent factors (Plasma KP, CSF KP, DMPFC-BG connectivity, and Anhedonia) within a single unified model, as illustrated in **Figure 4A**. A higher-order path model was used to control for high covariance between plasma and CSF KP metabolites. A combined KP factor including both CSF and Plasma KP (Cf(95%)=0.39(0.28, 0.58), t=5.1, p<0.001), plasma KP factor (Cf(95%)=0.27(0.20, 0.39), t=5.13, p<0.001), and CSF KP factor (Cf(95%)=0.15(0.01, 0.27), t=3.5, p<0.001) were all indirectly associated with Anhedonia. Intriguingly the path from plasma KP to CSF KP to combined KP factor to DMPFC-BG connectivity to Anhedonia was significant (Cf(95%)=0.1(0.05, 0.19), t=2.7, p=0.007). The analytic details and loading coefficients are detailed in Supplementary Tables 5-7.

**Figure 4:**
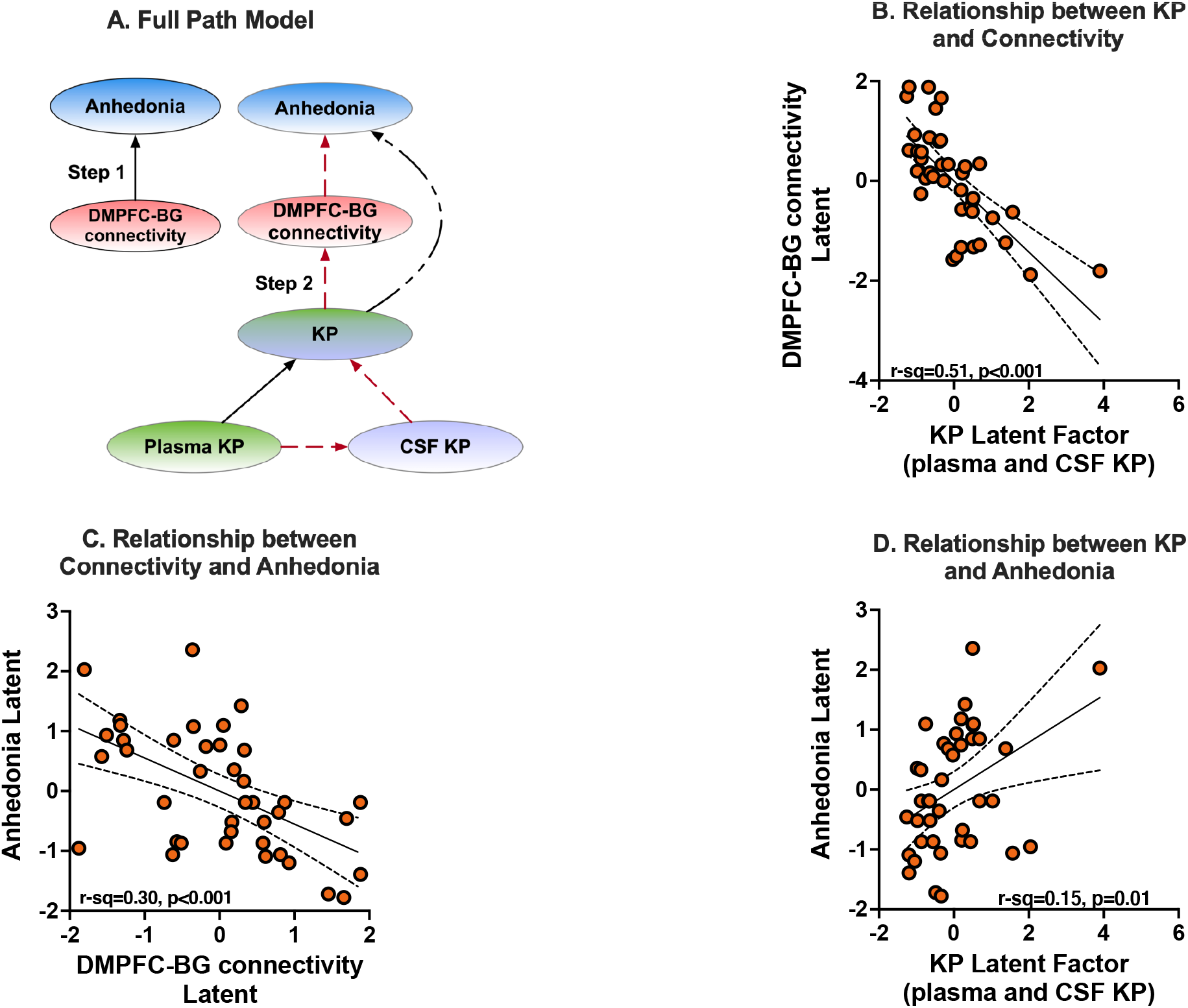
Connectivity-Anhedonia Associations: **Legend: Figure 4A** presents the overall scheme of the path model between all the latent factors within a single, reflective, multi-level path model. Briefly, kynurenine (KYN) and 3-hydroxy-kynurenine (3HK) in plasma and CSF were loaded onto their respective plasma and CSF kynurenine pathway (KP) factors that loaded further onto a combined KP factor. The KP factor was then related to dorsomedial prefrontal (DMPFC)-basal ganglia (BG) latent factor that was further linked to the Anhedonia latent factor described earlier. DMPFC-BG latent factor was composed of both plasma and CSF connectivities selected based on factor loadings (loading coefficient >0.7). 3/4 of the above connectivities, DMPFC-DMPFC (right to left DMPFC), DMPFC-left ventrolateral prefrontal region (LVLPFC), DMPFC-left dorsolateral prefrontal region (DLPFC), were correlated with plasma KYN, and 1/4, i.e., DMPFC-left basal ganglia (LBG) was correlated with CSF 3HK. The red-colored broken lines represent direct paths that combined to create an indirect or mediation effect. Step 1 (**Figure 4A**) examined a direct relationship between functional connectivity and Anhedonia. A significant negative association was noted between the DMPFC-BG connectivity factor and the Anhedonia factor (r-sq=0.34, p<0.001, not in the figure). **Figures 4B, 4C, and 4D** represent linear associations between the latent factor scores. **Figure 4B** represents the direct association between KP and DMPFC-BG connectivity, and Figure 4C presents the direct association between DMPFC-BG connectivity and Anhedonia. **Figure 4D** represents the indirect association between KP and Anhedonia mediated by DMPFC-BG connectivity. A mediation effect of the link between KP and Anhedonia by connectivity and the relationship between Plasma KP and Anhedonia by CSF KP and connectivity was significant (details in text).

### Associations between MRS metabolites and DMPFC-BG connectivity

The details of this analysis are presented in **Supplemental Information 6 and Supplementary Figure 4**.

### Power Analysis, Rigor, and Reproducibility

Detailed power estimates and analyses are provided in **Supplemental Figure 5**. Most of our effect sizes (noted in appropriate sections) were in the strong effect size range with Cohen’s f-sq values >0.25. With a sample size of 40, alpha of 0.05, and power of 0.8, effect sizes > 0.25 (for direct paths and linear models) and >0.2 (for mediation) were adequate to identify key effects. We have presented data on multiple resampled analyses and sensitivity analyses examining the impact of covariates and disruptive noise factors. The lasso-inferential models used 100-fold resampling of lasso coefficients, and PLS-SEM used 10000-fold bootstrapped CI.

## Discussion

Herein, we report that plasma QA, KYNA, and KYNA/QA were associated with MRS signals of Glu in LBG. Plasma KYN/TRP and CSF 3HK were associated with mI signals in the same region, although this association was subject to moderation by age and BMI. MRS Glu and mI are believed to index astroglial dysfunction and distress^57,58^. Astroglial dysfunction is a well-documented feature of mood disorders but may be particularly relevant to the inflammatory subtype of MD^59,60^. The precise nature of the astroglial dysfunction in the inflammatory subtype of depression is not yet fully clarified. It is now widely accepted that ^1^H-MRS primarily measures intracellular Glu that exists at much higher concentrations (5-15 mM) compared with the lower concentrations of Glu (~5-10 μM) encountered in the extracellular space^61^. A neuronal-astrocytic neurotransmitter cycle exists in the brain in which Glu from the neuronal pool is released into the synaptic cleft as a neurotransmitter, taken up by astrocytes, converted to glutamine, and returned to the neuron in this synaptically inactive form and converted back to glutamate^62,63^. Neuronal Glu processing by the astroglia is an energy-intensive process involving an expenditure of 2 molecules of adenosine triphosphate (ATP) for each molecule of glutamate processed^62^. Under mild anesthesia, Glu-glutamine neurotransmitter cycling is believed to consume >80% of total glucose oxidation in the cell^62^. However, the energy costs of glutamate neurotransmission need to be weighed against the energy demands induced by inflammation. For instance, when immune cells are stimulated in vitro, they consume ~25-30% more energy, which increases ~60% during acute infections^64^. Thus, MRS Glu build-up in inflammation may reflect the impaired processing capacity resulting from astrocytic distress. MRS Glu increases have been reported in various inflammatory and neuropsychiatric disorders^65^. Despite its presence in other tissues, mI is believed to be an astroglial marker as it is highly concentrated in the astroglial cells, as shown in vivo NMR s studies^57,58^. It participates in the osmoregulation of the astrocytic cell and is highly sensitive to inflammation and oxidative stress^57,58,66^. Increases in mI signals have been associated with immune pathologies such as multiple sclerosis, human immunodeficiency virus (HIV), and hepatitis C virus infections^66^. Intriguingly, it has also been strongly associated with aging and neurodegenerative dementia^66^.

KP metabolism may reflect the convergence of pathologies involving bioenergetic dysfunction, Glu-cycling, and astroglial distress. More recently, the transport of KP molecules across BBB via LAT-1 has been emphasized, as it may be targeted by leucine, a high-affinity blocker of KP transport across LAT-1^14^. Upon entry via LAT-1, KYN, and 3HK may bind to the nod-like receptor protein type 2 (NLRP2) in the astrocytes leading to amplification of neuroinflammatory response via activation of caspase-1 and nuclear factor kappa-B (Nf-Kb), culminating in the release of (IL)1b^17^. Thus, kynurenine may by itself act as a danger-associated molecular pattern (DAMP) that stimulates astrocytic inflammation resulting in increases MRS Glu and mI. Basal ganglia is a highly vascularized region with many end arteries offering a rich BBB interface for such pathological immune-KP-brain interactions. Increasing inflammation of the glial cells in the BG region may account for at least some of the reward circuit dysfunction and resultant anhedonia evident in this study.

KP was associated with impaired functional activity and connectivity within/between DMPFC-BG regions, which in turn was associated with anhedonic items of the IDS-SR. Optogenetic experiments indicate that both the top-down (DMPFC Glu systems) and bottom-up (brainstem dopamine systems) compete for functional dominance over the striatal regions to balance competing motivational demands^67^. While the bottom-up inputs convey prediction error reward signals, the top-down inputs adjust and reorient motivational priorities to facilitate longer-term or non-hedonic objectives. For instance, water (during dehydration) or rest (during inflammation and infection) may be preferred over more powerful, reinforcing hedonic stimuli such as sucrose water^68^. Impairments of DMPFC might identify a distinct subgroup of depressed patients who may respond to targeted stimulation of this region^69–71^. DMPFC and its downstream connections have been proposed as promising stimulation targets using transcranial magnetic stimulation (TMS) for anhedonic depression^72^. Our data build upon this developing literature by demonstrating that KP molecules specifically target the DMPFC-BG frontostriatal system.

A major thrust of this study is to focus attention on the anhedonic subtype of depression. Anhedonia refers to a decreased ability to enjoy previously pleasurable experiences experiences^73^. Anhedonia may be viewed as a transdiagnostic entity seen among many neuropsychiatric disorders^74–76^. Our data support conceptualizing anhedonia as a dysfunctional integration between top-down and bottom-up networks that regulate reward functions disrupted by inflammation and its downstream products such as KP molecules^68,77^. Anhedonia is not only common in late-life depression (rates of ~70%) but appeared to be associated with a ~30% increase in the risk of death and disability among these subjects^78^.

The study’s strengths include an in-depth characterization of KP molecules across plasma and CSF compartments, chemical and functional profiling of frontostriatal brain regions, and relating brain changes to anhedonia. However, there are limitations to consider. First, our sample lacks a control group, although we intended to examine KP dysfunction specifically among depressed subjects. Induction of anhedonia among healthy subjects by immune stimulation is well known to engage similar top-down and bottom-up brain circuits^79–82^. Second, our data is cross-sectional. Longitudinal or interventional studies will provide more insights into the above-noted interactions. Our sample is relatively small, though the effect sizes were strong enough to support the sample size in power analyses. The sample was drawn from and represents a limited universe of medication-free, depressed individuals who were willing to undergo MRI scanning and CSF sampling. Our focus on anhedonia may have excluded other depressive symptom dimensions of interest. Suicidality has been significantly related to KP metabolism over several studies. The recruitment criteria for the study excluded patients with significant suicidal ideation. Finally, given our focus on frontostriatal systems, we did not examine other brain networks.

In conclusion, the study’s findings indicate that KP metabolites among depressed subjects were associated with chemical and functional changes in key hub regions of the frontal-subcortical reward system, leading to symptoms of anhedonia. Targeting the peripheral KP system or its transfer into the brain are viable areas of therapeutic interest. Besides, the effect of KP metabolites on brain chemistry may be mitigated by Glu modulating agents such as ketamine and riluzole. Similarly, the newly emerging field of glial protective agents may help ameliorate the toxic effects of inflammation and KP metabolites on the brain.

## Data Availability

Not available.

## Table/Figure Legends

**Table 1: Demographic, Clinical and** Biological **Variables**: **Legend: Table 1** Abbreviations: BMI=body mass index, HAMD=17 item Hamilton Depression rating scale, LBG= left basal ganglia, Glu=glutamate, mI=myo-inositol, TNF=tumor necrosis factor, TNFR2=tumor necrosis factor receptor type 2, TRP=tryptophan, KYN=kynurenine, KYNA=kynurenic acid, AA=anthranilic acid, 3HK=3-hydoxy kynurenine, 3HAA=3-hydoxy anthranilic acid, QA=quinolinic acid, CSF=cerebrospinal fluid.

