## Supplementary material for "Kynurenines Increase MRS Metabolites in Basal Ganglia and Decrease Resting State Connectivity in Frontostriatal Reward Circuitry in Depression": F_Supplemental Information.pdf

#### Supplementary Information 1: Estimation of KP Metabolites

Estimation Parameters for KP Assays have been presented in our earlier papers [1]. Estimation details for **immune markers** have been provided as Supplements to our previously published papers[2-4]. All assays were conducted at Brains Online (now part of Charles River Labs, Inc). Concentrations of TRP, KYN, KA, 3HKYN, AA, 3HAA and QA in plasma and CSF were measured blind to diagnosis by Brains Online, LLC (Charles River, Inc, https://www.criver.com/consult-pi-ds-cns-research). The serum and CSF metabolite concentrations were estimated using high-performance liquid chromatography (HPLC) with tandem mass spectrometry (MS/MS) detection using standardized protocol per specifications provided by the vendor (see supplementary information for more details). The overall CV for the assays were <10% for TRP, KYN, KA, 3HKYN and QA. KYN/TRP (KT) and QA/KA (QK) ratios were computed in the plasma and CSF fractions for further analyses.

#### Supplementary Table 1: Estimation Parameters for Kynurenine-Pathway Metabolites

| Analysis Columns | Study Min | LLOQ | %CV |
|---|---|---|---|
| Plasma TRP (µM) | 35 | 3 | 2.3 |
| Plasma KYN (µM) | 0.91 | 0.225 | 1.84 |
| Plasma KYNA (nM) | 15.1 | 7.5 | 4.18 |
| Plasma AA (nM) | 3.08 | 3 | 6.46 |
| Plasma 3HK (nM) | 16.7 | 5 | 4.54 |
| Plasma 3HAA (nM) | 8.77 | 10 | 6.27 |
| Plasma QA (nM) | 226 | 100 | 4.89 |
| CSF TRP (µM) | 0.47 | 0.12 | 3.82 |
| CSF KYN (nM) | 13.7 | 12 | 4.22 |
| CSF KYNA (nM) | 0.71 | 0.6 | 3.56 |
| CSF AA (nM) | 1.49 | 1.2 | 3.82 |
| CSF 3HK (nM) | 1.27 | 1.2 | 4.59 |
| CSF 3HAA (nM) | excluded | 1.2 | 6.3 |
| CSF QA (nM) | 10.5 | 5 | 4.72 |

**Legend:** The first column specifies study minimum and second and third columns provide assay detection limits. * = CSF 3HAA was excluded as most of the assay values were unreliable.

**Abbreviations:** LLOQ = lower limits of quantification, %CV= coefficient of variability of the assay, TRP=tryptophan, KYN=kynurenine, KA=kynurenic acid, AA=anthranilic acid, 3HKYN=3-hydroxy kynurenine, 3HAA=3-hydroxy anthranilic acid, QA=quinolinic acid

#### Supplementary Information 2: MRI methods

**MRI data acquisition:** MRI scans were conducted using a Siemens 3T Trim-Trio scanner (Siemens Medical Solutions, Malvern, PA, USA). Foam restraints were used to limit head motion. **T1 images** were acquired using a magnetization prepared rapid gradient echo (MPRAGE) sequence (TR / TE / TI = 2300 / 3.02 / 1100 ms, flip angle (FA) = 8$^{o}$, voxel size = 1 x 1 x 1 $mm^3$, matrix size = 256 x 256). **Single-voxel magnetic resonance spectroscopy (MRS)** data were acquired using a point-resolved spectroscopy (PRESS) sequence with chemical shift selective (CHESS) pulses for water suppression (TR / TE = 3000 / 30 ms, FA = 90$^{o}$, sampling size = 1024, size of voxel of interest (VOI) = 30 x 17 x 17 $mm^3$). The VOI was placed in the left basal ganglia area. There were 2 scans per VOI with the same parameters: a scan of 4 averages without water suppression and a scan of 128 averages with water suppression. The spectrum peak width, measured as full width at half maximum (FWHM) of water signal, was controlled to be less than 15 Hz by adjusting VOI location and shimming before the scans. **Resting-state fMRI (rsfMRI) data** were acquired using a Z-saga echo planar imaging (EPI) [5] sequence (TR / TE1 / TE2 = 2950 / 30 / 67 ms, FA = 90$^{o}$, FOV = 220 mm, matrix size = 3.4 x 3.4 x 4.0 $mm^3$, slice number = 30, 150 repetitions). Participants were required to look at a cross sign on a screen during the scan.

**MRI data analysis:** FreeSurfer[6-8] and LCModel[9,10] were used to process structural MRI and MRS data analysis respectively. AFNI[11,12] was used for all other analyses if not specifically mentioned below. T1 Images were processed to generate gray matter (GM), white matter (WM) and ventricle+cerebrospinal fluid (CSF) segments with FreeSurfer. They were also used to register with and then convert the RS-fMRI images into the Montreal Neurological Institute (MNI) space.

**MRS-SVS data**: Details of processing steps have been provided in our previous publication[4]. Briefly, key processing steps included: 1) A VOI was generated on T1 images based on the VOI location and size. 2) Volumetric ratios of GM, WM, and CSF ($R_{GM}$, $R_{WM}$, $R_{CSF}$) were calculated for the VOI with the GM, WM and CSF segments generated on the T1 images. 3) Water concentration (WCONC) of the VOI was calculated with the formula: WCONC = (43300 x $R_{GM}$ + 35880 x $R_{WM}$ + 55556 x $R_{CSF}$) / (1 - $R_{CSF}$), as specified in LCModel manual[10]. 4) Applied the WCONC in LCModel for partial-volume correction of absolute metabolite concentrations, which were estimated using water-scaling method. 5) Concentration data with SD%<20% (SD%: estimated standard deviations, Cramer-Rao lower bounds, expressed in percent of the estimated concentrations) were used for further analysis.

**Resting State fMRI** analysis have been described extensively in our previous publication[13]. Key preprocessing steps included: removal of signal spikes, slice-timing shift correction, motion correction, and removal of motion and CSF signal with a general linear model (GLM). The residual time-series data were used to calculate regional homogeneity (ReHo) value for each voxel. The ReHo maps were converted into the MNI space and spatially smoothed with FWHM = 4 mm for the ReHo-KP (plasma KYN, plasma 3HK, plasma KYN/TRP ratio, CSF KYN, and CSF 3HK) correlation analysis across subjects. Regions of interest (ROIs) identified within these correlation maps, were tested for cluster-wise multiple-test correction with voxel-level p <= 0.001 and cluster-wise p <= 0.05[14,15]. Mean ReHo of the ROIs were calculated and used for further statistical analysis. To identify brain regions connected to the ReHo ROIs, seed-based connectivity analysis was performed following these steps: 1) Removal of the WM signals, temporally filtering (0.01–0.1 Hz) and spatial registration to the MNI space for the above preprocessed time series data for each subject. 2) Averaged time series data were calculated for the individual ReHo-ROIs, which were then used for voxel-wise, seed-to-whole brain correlation analysis. The correlation data were converted to z-score data. The seed-connectivity target ROIs whose z-score data were correlated with KP measures were identified for the CSF kynurenine (CSF KYN and 3HK) and plasma kynurenine (plasma KYN, 3HK, KYN/TRP ratio) separately. Thus, the z-score data generated from CSF-KP seed (the ReHo ROIs defined with the CSF kynurenine measures) were correlated with the CSF kynurenine measures, and those from the plasma-KP seed were correlated with the plasma kynurenine measures, respectively. The target ROIs (CSF-KP or plasma-KP targets) were identified with the same cluster-wise multiple-test correction method mentioned above. Averaged z-score data were calculated for these target ROIs for further statistical analysis.

### Supplementary Information 3: Statistical Analysis Plan

***Lasso inferential models*** were used to identify relationships between KP markers and MRS metabolites in the basal ganglia region. Independent models were used to predict each of the key metabolites available from MRS, including glutamate, myo-inositol, choline, creatine, and NAA. Similarly, independent models were used for plasma and CSF KP, with age, sex, race, and BMI being included as control factors common to all models. The lasso-inferential algorithm partials out the effects of covariates before using the relationship between predictors of interest and response variables. The partialing out method uses previously validated techniques that have long been used for ordinary least squares approaches. This approach provides easily interpretable linear estimates, including parameters estimate, standard error, and z-values that would yield significant values. None of these are available in the traditional lasso selection methods. Lasso inferential models were conducted in Stata-version 16. The models were conducted in three phases. We used the default "plug-in" (heteroscedastic) to identify primary variables of interest in the first phase. The plug-in method is relatively insensitive to noise and is very useful for identifying key effects of interest. The second phase consisted of the plug-in method combined with k(10)-fold x resampling (#10 reps), totaling 100 repetitions. We used this method to examine the predictive stability of the estimates over repeated resampling with variable k-fold repetitions. The third phase consisted of sensitivity analysis using alternative validation methods, including cross-validation (CV), adaptive, and minimum Bayesian Information Criteria (BIC)-based approaches. The sensitivity analysis examined if the relationships identified by the primary plug-in model would be disrupted by adding noise factors to the model. For instance, the CV approach includes many control covariates, and the adaptive method uses a relatively lesser number of control covariates than CV - but more than the plug-in. The BIC is manually selected and thus may have a variable number of control covariates. Thus, sensitivity analysis helps to differentiate if increasing or decreasing the number of control covariates would alter the stability of the primary findings.

***Rationale:*** The data points provide two major difficulties. There are many covariates (high-dimensional models), and many of them are strongly correlated with each other. High-dimensional models have a few covariates of interest and many potential control covariates. The number of potential covariates can be larger than the sample size. We want to estimate the coefficients on the few covariates of interest. High-dimensional models force us to make covariate selection. The assumption that the number of covariates in the model is small relative to the number of observations in the data is called a 'sparsity assumption.' Correlations between predictors can be a major source of prediction error but leading to variance inflation.

***Lasso-inferential Models:*** To overcome these factors, we used the least absolute selection and shrinkage operator (lasso) to make covariate selection. The lasso does better covariate selection than non-data-based methods. Lasso models use covariate selection by penalizing coefficients that are subject to variable inflation. However, the lasso models still make some small mistakes in which covariates to include and exclude. In addition, the models do not help in inference as the coefficients generated do not satisfy linear moment conditions. Lasso-inferential models (Stata v16) use the lasso for covariate selection but estimate the effect of interest using moment conditions robust to the mistakes the lasso makes.

***Cross-fit, Partialing out (XPO) estimator:*** This is accomplished by partialing out (PO) estimators. PO estimation is an extension of the partialing-out method for obtaining the ordinary least squares (OLS) estimate for the coefficient and standard error on d (also known as the result of the Frisch-Waugh-Lovell theorem). Cross-fitting is also known as double machine learning (DML), uses split-sample techniques on PO estimators to weaken the condition that restricts the number of covariates in the model. Mechanically, split-sample techniques split the sample into K subsamples called folds. The lasso estimators use the data, not in fold k, to get (post-selection) coefficient estimates for each fold k, then use coefficients computed in step 1 to fill in residuals for the next analysis step.

***Tuning Parameters:*** The tuning parameters determine which covariates are included in the lasso and set before using the lasso for prediction or model selection. Plug-in methods, cross-validation, and the adaptive lasso are used to select the tuning parameters. Plug-in methods were used as the primary methods for variable selection. Plug-in estimators find the value of the λ that is large enough to dominate the estimation noise and tends to include the important covariates and excluding covariates that do not belong in the model. Cross-validation (CV) selects the λ value that minimizes the out-of-sample mean squared error (MSE) of the predictions and is excellent at including the important covariates. However, a CV includes many additional covariates that do not belong in the model. The adaptive lasso is a multistep version of CV. It is excellent at including the important covariates and, but it tends to include some extra covariates that do not belong in the model.

***Sensitivity Analysis:*** Including too many additional covariates can cause any of our robust estimators to perform poorly and slows the convergence rate of cross-fit lasso estimators. Sensitivity analysis uses diagnostics to determine how sensitive is the response of the main candidate estimator to small changes in model specification. We used the primary plug-in method to identify the main candidate estimator, used CV, the adaptive lasso, and hand-specified λ for sensitivity analysis.

**Path analysis** was conducted using consistent partial least squares -structural equation modeling (PLSc-SEM) in Smart PLS[16].

***Measurement Model:*** A formative measurement model was used based on our interest in examining the brain effects of KP markers that readily crossed the BBB (KYN and 3HK). Similarly, all DMPFC-BG related connectivities (total=4) were tested. Based on previously published recommendations[17], formative measurement models were used after examining convergent validity, indicator collinearity, and indicators' statistical relevance to the underlying construct. The robust correlations between KP markers and their constructs established convergent validity to their constituent indicators (KYN, 3HK) in each of the compartments (plasma, CSF) as reported in our earlier publication[1]. The underlying dimensions of connectivity were identified based on their correlation with KP markers, thus establishing a full convergent validity for the tested constructs. The variance inflation factor (VIF) was used to evaluate the collinearity between formative indicators. We maintained all VIF values at <4 to accommodate seeds and target regions located close to each other, though identified as different regions of interest in the voxel-based analysis. The indicator's contribution to the overall construct was measured by its outer loading coefficient (i.e., the bivariate correlation between the indicator and the construct being >0.7) and its beta weights. The outer loading coefficient in PLSc-SEM is akin to the communality coefficient in common factor analysis. Based on formative measurement theory, all indicators loading more than the 0.7 value were retained irrespective of the significance of their beta weights in the regression equation. This step is necessary to capture individual contributions of all formative indicators to the entire domain of the construct, irrespective of their significance levels.

***Structural Model:*** Assessment of structural model was based on relevant associations between various latent constructs (i.e., direct effects). Significance testing of path coefficients was performed using 10000-fold, bias-corrected bootstrapping. The structural model was assessed using the coefficient of determination (r-sq). The overall effect size of the individual values was established using Cohen's f-sq values to establish the model differences with and without the given variable of interest. We also obtained similar effect size estimates for **all effects** in each model by comparing it against a null or intercept-only model. However, r-sq only indicates the model's in-sample explanatory power without providing much information on the model's out-of-sample predictive power. Out-of-sample prediction involves estimating the model on an analysis (i.e., training) sample and evaluating its predictive performance on data other than the analysis sample, referred to as a holdout or validation sample. We used blind folding-based cross-validated redundancy measures (Q-sq) to measure out-of-sample prediction power. The Q-sq values are parallel to r-sq values and can be used to calculate q-sq, scaled similarly to Cohen's f-sq for r-sq values. Indirect effects on a given target construct by one or more intervening constructs were tested using assessment of **mediating effects**. We also computed total effect, defined as the sum of the direct and all indirect effects, to measure the directionality of mediating effects (full versus partial mediation). The approach for testing mediation effects were conducted using steps outlined by Baron and Kenny[18].

#### Supplementary Information 4: Sample and Recruitment

Forty-nine subjects with available measures of KP and MRS measures participated in the study. 3/49 subjects (ID# 52,105,112) had at least 1 missing values of plasma KP metabolite (one subject with missing KYNA and two subjects with missing HAA and AA). Thus, 46 subjects had all 7 plasma KP measures. Although 41 had resting state measures, only 40 of them had CSF KP measures of interest (i.e., plasma, CSF KYN and 3HKYN). Thus 46 subjects with full of plasma KP and MRS measures, 40 of whom had CSF KP and resting fMRI measures were included in the study. 7/49 subjects did not undergo CSF sampling, and 11 other subjects had missing individual CSF KP markers. Individual KP marker missingness was attributed to <lower limits of quantitation (described in detail in our earlier publication). No discernible group differences in background variables (age, sex, race, BMI, depression severity) were noted between the groups with (n=31) and without (n=49) the full panel of plasma/CSF markers. As reported in our earlier publications, high correlations were noted between KP markers in the plasma and CSF (Bonferroni-corrected $p<0.05$). The values of plasma and CSF KP markers were log-transformed for normalization. Missing plasma and CSF KP measures were not imputed as the data was not missing at random. 1/49 subjects did not provide information in response to item #22 on sexual function in depression. The precise reason was not known, but the missingness may have been at random. Consequently, this value was imputed using mean value replacement in the path modeling software.

#### Supplementary Table 2: Missing Data Information

| Study Measure | N | N Missing | N Available | %Missing |
|---|---|---|---|---|
| Plasma TRP (µM) | 49 | 0 | 49 | 0 |
| Plasma KYN (µM) | 49 | 0 | 49 | 0 |
| Plasma KYNA (nM) | 49 | 1 | 48 | 2 |
| Plasma AA (nM) | 49 | 2 | 47 | 4 |
| Plasma 3HKYN (nM) | 49 | 0 | 49 | 0 |
| Plasma 3HAA (nM) | 49 | 2 | 47 | 4 |
| Plasma QA (nM) | 49 | 0 | 49 | 0 |
| CSF TRP (µM) | 49 | 8 | 41 | 16 |
| CSF KYN (nM) | 49 | 8 | 41 | 16 |
| CSF KYNA (nM) | 49 | 13 | 36 | 27 |
| CSF AA (nM) | 49 | 7 | 42 | 14 |
| CSF 3HKYN (nM) | 49 | 7 | 42 | 14 |
| CSF QA (nM) | 49 | 12 | 37 | 24 |
| MRS data | 49 | 0 | 49 | 0 |
| ReHo data | 49 | 9 | 40 | 18 |
| Connectivity data | 49 | 9 | 40 | 18 |
| Behavior | 49 | 1 incomplete item on IDS-SR* | 49 | 0 |
| Plasma KP/MRS | 49 | 3 | 46 | 6 |
| CSF KP/Resting State Data | 49 | 9 | 40 | 18 |

*= 1 subject did not to provide information on item #22 on Libido in IDS-SR.

#### Supplementary Information 5: MRI results

Brain regions in which ReHo were correlated with plasma (KYN and 3HK) or CSF (KYN and 3HK) KP measures are shown as the blue areas in **Supplementary Figure 1**. These regions of interest (ROIs) are referred to as plasma-KP and CSF-KP ROIs, respectively. The plasma-KP ROIs included the right OC (R-OC), right DLPFC (R-DLPFC), left DMPFC (L-DMPFC), bilateral BG (B-BG), right DMPFC/ACC (R-DMPFC/ACC), bilateral DMPFC (B-DMPFC), and right prefrontal white matter (not shown in Supplementary Figure 1). The CSF-KP ROIs included the R-DLPFC, left BG (L-BG), right DMPFC/MCC (R-DMPFC/MCC), B-DMPFC, and left cerebellum (not shown in Supplementary Figure 1).

The ReHo ROIs were also set as the plasma-KP or CSF-KP seeds (***denoted with the red or green circles in Supplementary Figure 1***) to identify brain regions in which the seed-connectivity z-score data were correlated with the plasma and CSF kynurenine measures, respectively. These brain regions are referred to as seed-connectivity target ROIs (***underscores represent connectivity***). As shown in Supplementary Figure 1, the plasma kynurenine-related connections (plasma-KP Links [red circle_red square], 12 in total) included: B-DMPFC_ L-VLPFC, B-DMPFC_ L-DLPFC, B-DMPFC_ B-DMPFC, B-DMPFC_ B-thalamus, B-DMPFC_ R-VLPFC, L-DMPFC_L-PSSJ, L-DMPFC_L-IPC, L-DMPFC_L-precuneus, R-DMPFC/ACC_L-VLPFC, R-DMPFC/ACC_L-DLPFC, R-DMPFC/ACC_B-thalamus, and B-BG_L-LOFC. The CSF kynurenine-related connections (CSF-KP Links [green circle_green square], 7 in total) included: B-DMPFC_L-BG, B-DMPFC_R-BG, R-DLPFC_L-DLPFC, R-DMPFC/MCC_L-OC, L-BG_L-BG, L-BG_R-BG, and L-BG_L-DMPFC. The coordinates of the ReHo / seed ROIs and seed-connectivity target ROIs are listed in **Supplementary Tables 1, 2**.

#### Supplementary Table 3: KP-Correlated ReHo ROIs

| ROIs | | Coordinates for the Center of Mass | | |
|---|---|---|---|---|
| | | x (mm) | y (mm) | z (mm) |
| **Plasma-kynurenine pathway (Plasma-KP)** | | | | |
| ReHo-Plasma 3HK | R-DMPFC/ACC | -3.3 | -45.4 | 27.9 |
| | B-BG | -3.6 | -11.7 | -2.3 |
| | R-OC | -33.7 | 93.8 | 1.8 |
| | L-DMPFC | 12.0 | -41.4 | 53.4 |
| ReHo-Plasma KYN | B-DMPFC | 5.5 | -47.7 | 16.1 |
| | R-PFWM | -16.2 | -51.0 | 19.5 |
| | R-DLPFC | -24.9 | -34.7 | 40.2 |
| **CSF-kynurenine pathway (CSF-KP)** | | | | |
| ReHo-CSF 3HK | L-BG | 13.8 | -3.2 | 8.3 |
| | R-DMPFC/MCC | -8.9 | -34.7 | 48.4 |
| | L-cerebellum | 30.4 | 77.7 | -23.4 |
| ReHo-CSF KYN | B-DMPFC | -3.0 | -53.0 | 15.8 |
| | R-DLPFC | -39.9 | -49.0 | 18.2 |

**Legend:** 12 ReHo ROIs, expansion of abbreviations with Supplementary Figure 1 Legend
**Abbreviations:** 3HK: 3-hydoxy kynurenine, B-: bilateral, R-: Right, L-: Left, ACC: anterior cingulate cortex, BG: basal ganglia, CSF: cerebrospinal fluid, DMPFC: dorsomedial prefrontal cortex, DLPFC: dorsolateral prefrontal cortex, MCC: middle cingulate cortex, PFWM: prefrontal white matter, KP: kynurenine pathway, KYN: Kynurenine, OC: occipital cortex.

### Supplementary Table 4: Kynurenine-related seed-connectivity ROIs

| ROIs | | Coordinates for the Center of Mass | | |
|---|---|---|---|---|
| | | x (mm) | y (mm) | z (mm) |
| **Plasma-kynurenine pathway (Plasma-KP)** | | | | |
| Plasma 3HK-ReHo/seed | Plasma 3HK -target | | | |
| L-DMPFC | L-Precuneus | 4.4 | 55.6 | 18.8 |
| | L-IPC | 46.9 | 68.3 | 38.3 |
| | L-PSSJ | 58.6 | 24.9 | 13.7 |
| R-DMPFC/ACC | B-Thalamus | -1.4 | 13.2 | 8.5 |
| Plasma 3HK -ReHo/seed | Plasma KYN -target | | | |
| B-BG | L-LOFC | 28.0 | -30.2 | -14.5 |
| Plasma 3HK -ReHo/seed | Plasma KYN/TRP-target | | | |
| R-DMPFC/ACC | L-DLPFC | 24.4 | -30.8 | 49.3 |
| | L-VLPFC | 50.3 | -30.5 | -10.1 |
| Plasma KYN-ReHo/seed | Plasma 3HK -target | | | |
| B-DMPFC | R-VLPFC | -50.0 | -31.4 | -5.0 |
| | B-Thalamus | -0.4 | 12.1 | 8.9 |
| Plasma KYN-ReHo/seed | Plasma KYN/TRP-target | | | |
| B-DMPFC | B-DMPFC | -2.0 | -52.6 | 12.8 |
| | L-DLPFC | 25.5 | -28.6 | 52.3 |
| | L-VLPFC | 51.0 | -31.6 | -9.2 |
| **CSF-kynurenine pathway (CSF-KP)** | | | | |
| CSF 3HK-ReHo/seed | CSF 3HK-target | | | |
| L-BG | R-BG | -20.5 | 3.4 | 13.7 |
| | L-BG | 22.0 | -7.0 | 11.3 |
| | L-DMPFC | 5.8 | -34.3 | 26.6 |
| R-DMPFC/MCC | L-PVC | 14.3 | 105.8 | -3.8 |
| CSF KYN-ReHo/seed | CSF 3HK-target | | | |
| B-DMPFC | R-BG | -22.1 | -10.4 | 2.4 |
| | L-BG | 25.5 | -1.2 | 9.4 |
| R-DLPFC | L-DLPFC | 29.1 | -53.4 | 20.5 |

**Legend:** Seed-connectivity as a function of plasma and CSF KP. **Abbreviations:** 3HK: 3-hydoxy kynurenine, B-: bilateral, R-: Right, L-: Left, ACC: anterior cingulate cortex, BG: basal ganglia, CSF: cerebrospinal fluid, DMPFC: dorsomedial prefrontal cortex, DLPFC: dorsolateral prefrontal cortex, IPC: inferior parietal cortex, KP: kynurenine pathway, KYN: Kynurenine, KYN/TRP: Kynurenine/Tryptophan ratio, LOFC: lateral orbitofrontal cortex, MCC: middle cingulate cortex, OC: occipital cortex, PSSJ: postcentral, supramarginal and superior temporal junction, PVC: primary visual cortex, RH: right hemisphere, VLPFC: ventrolateral prefrontal cortex.

#### Supplementary Figure 1: ReHo Regions of Interest (ROIs) and Seed Connectivity

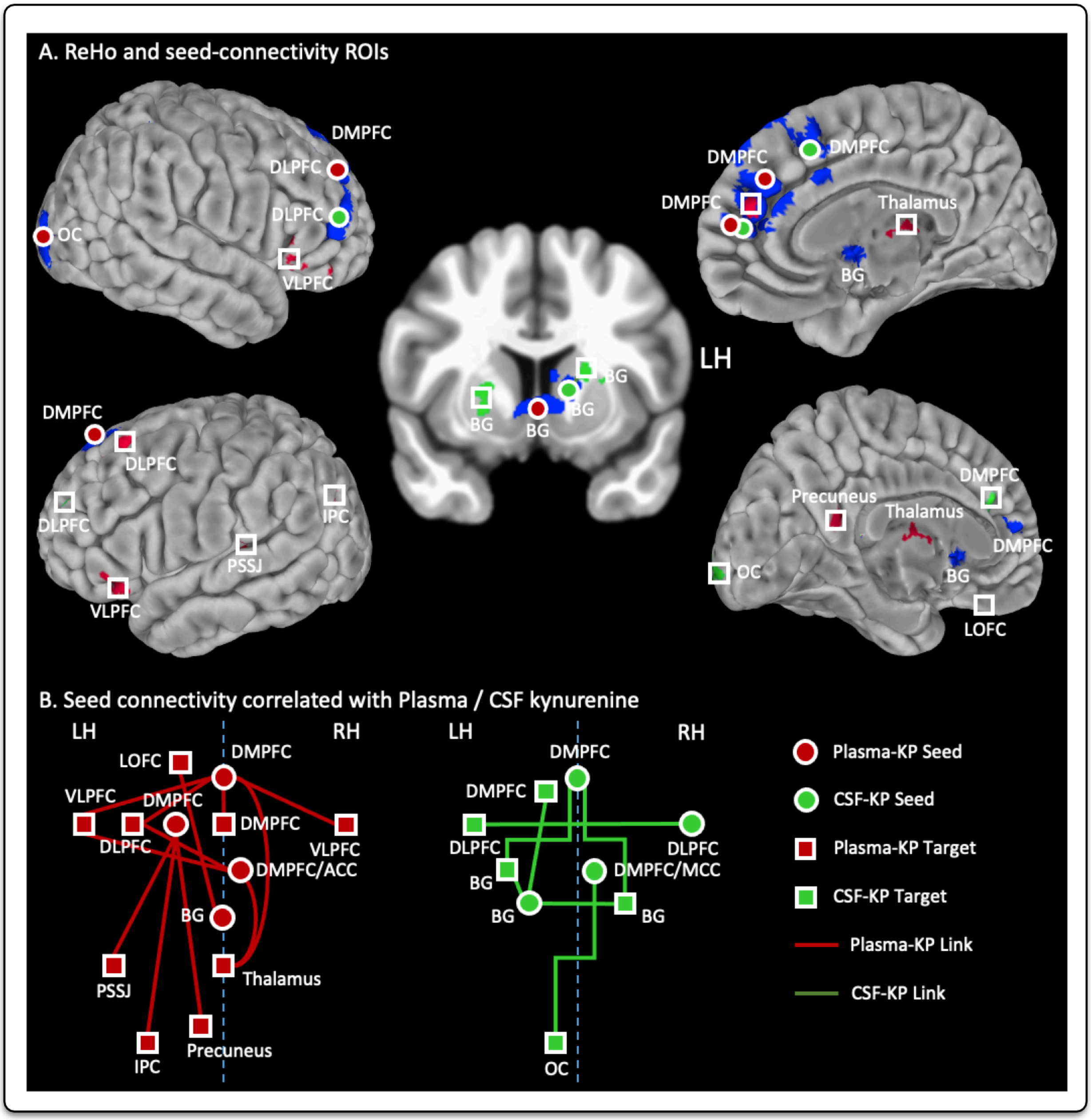

**Legend to Supplementary Figure 1.** ReHo and seed-connectivity ROIs correlated with the plasma (or CSF KP measures**. A. ROIs on brain images.** The blue areas show the combined plasma-KP and CSF-KP ReHo ROIs, in which ReHo values were correlated with plasma or CSF kynurenine measures. The plasma-KP ReHo ROIs are denoted with red circles, the CSF-KP ReHo ROIs are denoted with green circles**. B.** Schematic plot of the seed-connectivity ROIs. **Red lines:** connectivity correlated with the plasma kynurenine measures (Plasma-KP Link)**. Green lines:** connectivity correlated with the CSF kynurenine measures (CSF-KP Link). **Abbreviations:** 3HK: 3-hydoxy kynurenine, B-: bilateral, R-: Right, L-: Left, ACC: anterior cingulate cortex, BG: basal ganglia, CSF: cerebrospinal fluid, DMPFC: dorsomedial prefrontal cortex, DLPFC: dorsolateral prefrontal cortex, IPC: inferior parietal cortex, KP: kynurenine pathway, KYN: Kynurenine, KYN/TRP: Kynurenine/Tryptophan ratio, LH: left hemisphere, LOFC: lateral orbitofrontal cortex, , MCC: middle cingulate cortex, OC: occipital cortex, PSSJ: postcentral, supramarginal and superior temporal junction, PVC: primary visual cortex, RH: right hemisphere, VLPFC: ventrolateral prefrontal cortex

#### Supplementary Figure 2: KYNA/QA correlated with same Regions identified by KYN

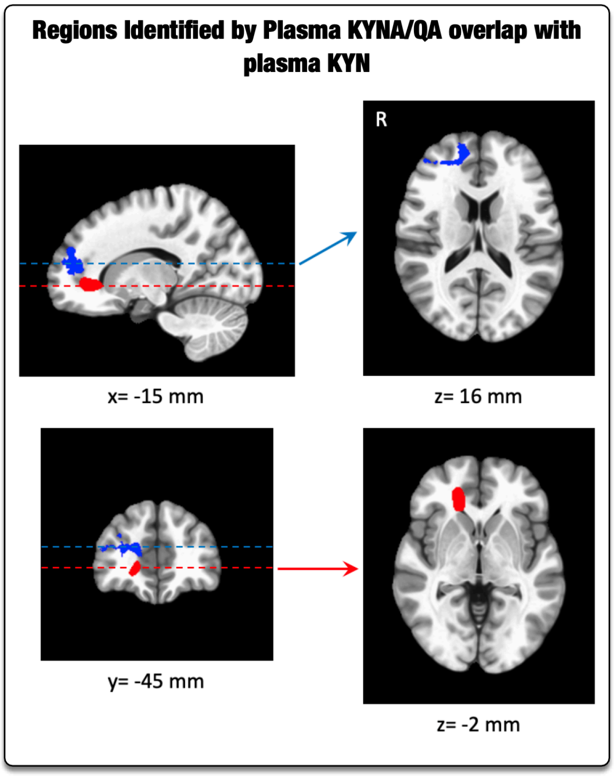

**Legend:** ReHo ROI correlates Plasma KYNA/QA and KYN in the same regions but in opposite directions.
Right prefrontal WM ROIs demonstrated that ReHo was associated with plasma KYNA/QA ratio (red area, center of mass coordinates: -17.6, -37.8, -3.1, **positive correlation**) and plasma KYN (blue area, center of mass coordinates: -16.2, -51.0, 19.5, **negative correlation**). The blue and red dashed lines show the locations of the axial slices on the right column. The x, y and z values are the coordinates in the MNI space. **Abbreviations:** R=right hemisphere, WM=white matter region, KYNA/QA=kynurenic acid/quinolinic acid ratio, KYN=kynurenine

#### Supplementary Figure 3: Plasma KYNA and QA Identify the Same Regions Identified by Plasma KYN/3HK

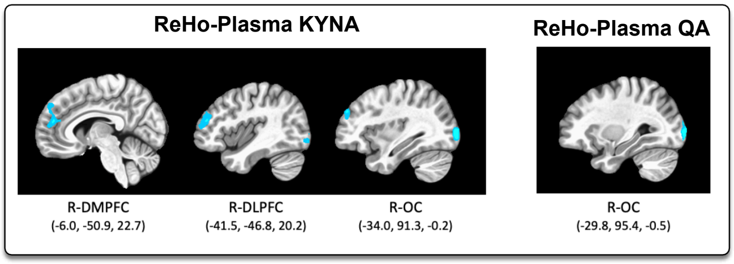

**Legend:** Compared to Supplemental Figure 1, Plasma KYNA and QA identified regions that were identical to those identified by plasma KYN and 3HK.

#### Supplementary Table 5: Path Analysis Indicator Loadings

| Indicator | Latent | Mean (SD) | t, p-value |
|---|---|---|---|
| #17. View of My Future | Anh | 0.778 (0.132) | 5.883, <0.001 |
| #19. General Interest | Anh | 0.788 (0.096) | 8.238, <0.001 |
| #22. Interest in Sex | Anh | 0.815 (0.077) | 10.648, <0.001 |
| #8. Response to Good/Desired Events | Anh | 0.641 (0.142) | 4.524, <0.001 |
| DMPFC1-LBG | Conn | 0.58 (0.17) | 3.401, 0.001 |
| DMPFC2-DMPFC | Conn | 0.825 (0.052) | 15.913, <0.001 |
| DMPFC2-LVLPVC | Conn | 0.751 (0.091) | 8.238, <0.001 |
| DMPFC3-LDLPFC | Conn | 0.815 (0.07) | 11.643, <0.001 |
| CSF 3HK (nM) | CSF | 0.696 (0.335) | 2.08, 0.038 |
| CSF KYN (nM) | CSF | 0.76 (0.119) | 6.393, <0.001 |
| Plasma 3HK (nM) | Plasma | 0.928 (0.035) | 26.772, <0.001 |
| Plasma KYN (µM) | Plasma | 0.916 (0.04) | 22.902, <0.001 |
| CSF 3HK (nM) | KP | 0.574 (0.273) | 2.098, 0.036 |
| CSF KYN (nM) | KP | 0.659 (0.097) | 6.816, <0.001 |
| Plasma 3HK (nM) | KP | 0.898 (0.055) | 16.179, <0.001 |
| Plasma KYN (µM) | KP | 0.867 (0.052) | 16.8, <0.001 |

**Legend:** Path Analytic Model Loadings. Dorsomedial prefrontal cortex (DMPFC), left basal ganglia (LBG), left ventrolateral prefrontal regions (LVPLC) and left dorsolateral prefrontal region (LDLPFC) are emphasized. DMPFC1,2,3 indicate regions of interest (ROI)s located in proximity and correlated with individual markers.

#### Supplementary Table 6: Direct, Indirect and Total Effects of Path Model

| Effects | Total | Direct | Indirect | %Direct |
|---|---|---|---|---|
| **CSF -> Anh** | 0.15 | | 0.15 | 100 |
| **Conn -> Anh** | -0.552 | -0.552 | | 0 |
| **KP -> Anh** | 0.393 | | 0.393 | 100 |
| **Plasma -> Anh** | 0.376 | | 0.376 | 100 |
| **CSF -> Conn** | -0.272 | | -0.272 | 100 |
| **KP -> Conn** | -0.712 | -0.712 | | 0 |
| **Plasma -> Conn** | -0.682 | | -0.682 | 100 |
| **Plasma -> CSF** | 0.66 | 0.66 | | 0 |
| **CSF -> KP** | 0.382 | 0.382 | | 0 |
| **Plasma -> KP** | 0.957 | 0.705 | 0.252 | 26.3 |

Legend: Relationships between latent factors representing CSF and plasma KP with DMPFC-BG connectivity (Conn) and Anhedonia (Anh), as described in Supplementary Table above.

#### Supplementary Table 7: Indirect/Mediation Effects of Path Model

| | Mean (SD) | t | P Values |
|---|---|---|---|
| **Plasma -> KP -> Conn -> Anh** | 0.277(0.05) | 5.56 | 0 |
| **Plasma -> CSF -> KP -> Conn -> Anh** | 0.099(0.037) | 2.7 | 0.007 |
| **CSF -> KP -> Conn** | -0.272(0.044) | 6.25 | 0 |
| **KP -> Conn -> Anh** | 0.393(0.077) | 5.09 | 0 |
| **Plasma -> CSF -> KP -> Conn** | -0.18(0.047) | 3.8 | 0 |
| **Plasma -> CSF -> KP** | 0.252(0.057) | 4.47 | 0 |
| **Plasma -> KP -> Conn** | -0.502(0.049) | 10.3 | 0 |
| **CSF -> KP -> Conn -> Anh** | 0.15(0.043) | 3.54 | 0 |

**Legend:** Mediational relationship between various latent factors in the structural model.

#### Supplementary Information 6: MRS-Connectivity Associations

We used **Self Validating Ensemble Modeling** (SVEM) to examine links between MRS markers age, sex, race, and BMI and DMPFC-BG connectivity (SVEM Module for JMP Pro v15, Predictum, Inc, Phoenix, AZ, USA). SVEM uses predictive ensemble modeling with fractionally-weighted bootstrapping methods customized for controlled, designed experiments using small samples[19,20]. The bootstrap method enables resampling the lasso coefficients over 3000 times. Variable importance is determined by the number of times non-zero lasso coefficients entered the equation against a random null variable added to the model. Maximum inclusion probability (MIP) represents the increase in selection probability normalized to the null (actual selection-null)/null.

**MRS-DMPFC-BG connectivity:** DMPFC-BG connectivity was entered as the response variable and all LBG MRS variables and covariates (main effects, 2-way interactions and quadratic effects) were entered as predictors into a supersaturated design into the SVEM modeling platform. Model validation metrics were remarkably similar in the training (r-sq=0.32, root average squared error, RASE=0.09) and holdout sets (r-sq=0.21, RASE=0.09), and the actual-by-predicted values of the response in the holdout set was significant (F (1, 18)=9.83, p=0.006, **Figure 4A**). Lasso-variable selection algorithm (**Figure 4B**) identified following significant effects [expressed as selection probabilities = mean(log odds)]: glutamate*inositol (1.97), age*glutamate (0.84), inositol (main effect, 0.75), BMI*glutamate (0.65), age*inositol (0.62), age*BMI (0.604), and glutamate^2 (quadratic effect, 0.59). Intriguingly, all these associations were negative, as indicated by the model-averaged coefficients.

### Supplementary Figure 4: MRS-Connectivity Associations

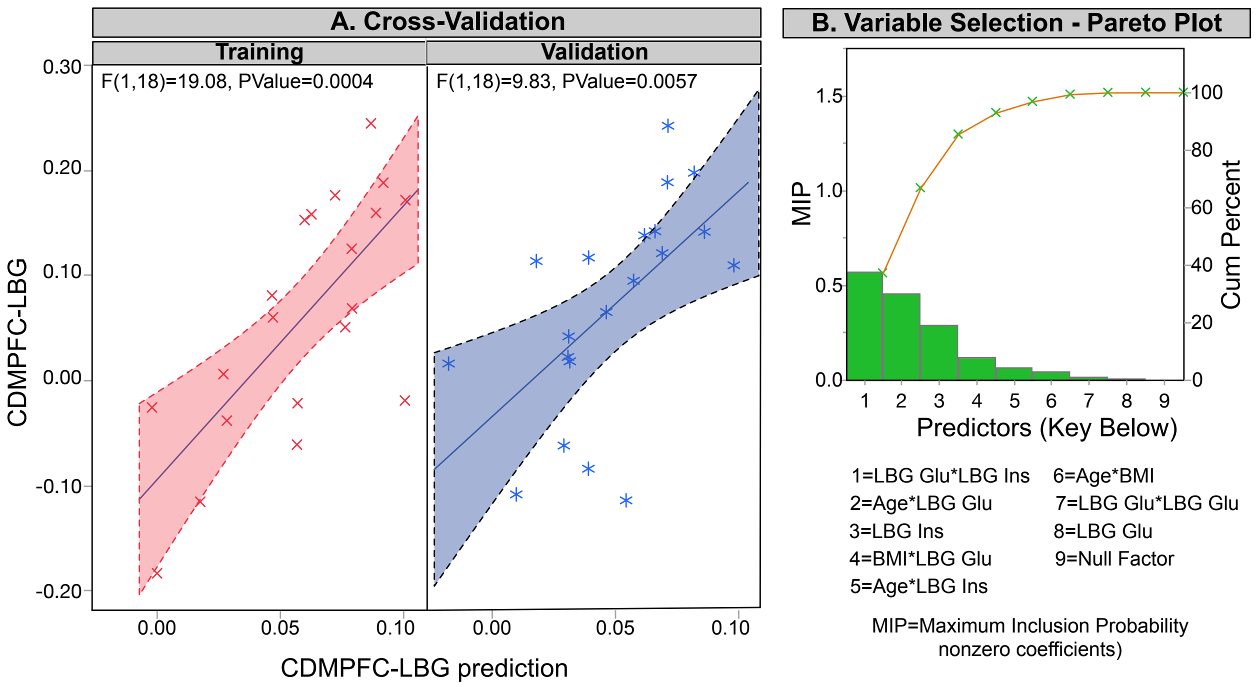

#### Supplementary Figure 5: Power Analysis Curve

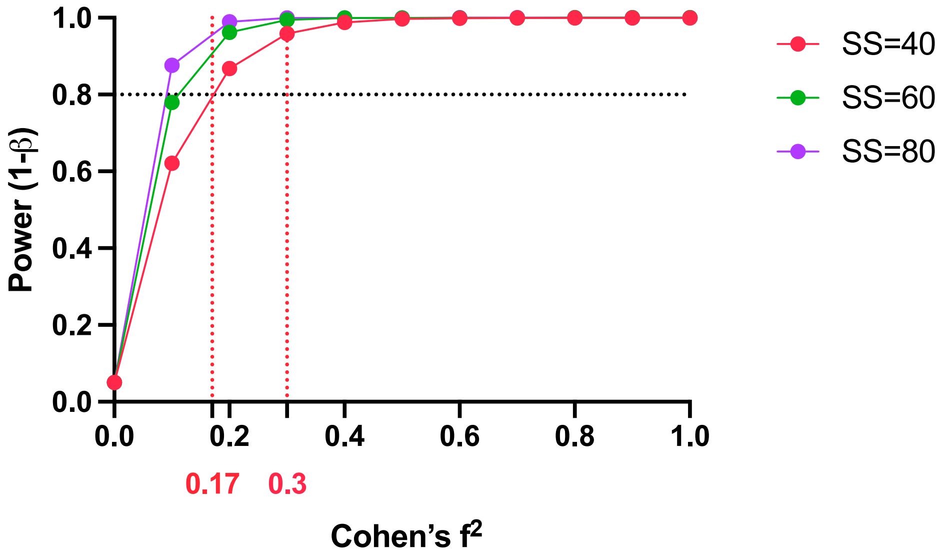

**Legend:** The figure plots represent the relationship between effect size estimates and power analysis for MRS and path analysis. The study effect sizes for MRS and power analysis are presented in red colored fonts on the x-axis. Effect sizes are represented in terms of Cohen's $f^2$ statistic. MRS analysis of left basal glutamate yielded an effect size of 0.17 that yielded a slightly suboptimal power of 0.70 for the sample size of 48 subjects with available for plasma and MRS glutamate measures). On the other hand, the path analysis of ReHo and seed-connectivity with Anhedonia indicated much stronger effect sizes >0.3 yielding power estimates >0.8, even with a sample size of 40.
